# Trends in Covid-19 hospital mortality in women and men

**DOI:** 10.1101/2021.09.06.21263166

**Authors:** Luis Ayerbe, Carlos Risco-Risco, Diego Martínez-Urbistondo, María Elena Caro-Tinoco, Salma Ayis

**Affiliations:** Centre of Primary Care. Queen Mary University of London. London, UK; Carnarvon Medical Centre. Southend on Sea. UK; Internal Medicine Department, Hospital Universitario HM Sanchinarro, Madrid, Spain; School of Population Health and Environmental Sciences, King’s College London, London, UK

## Abstract

**Introduction:** It remains unclear if the development of health services, clinical management, and scientific evidence, during the pandemic is associated with better medical outcomes, sustained in the long term, for Covid-19 patients of each gender. This study presents the trends in mortality associated with Covid-19 for women and men during the first year of the pandemic.

**Methods:** This study was based in 17 Spanish hospitals. Sociodemographic, clinical, and mortality data from all patients with Covid-19, who had been discharged alive, or had died after being admitted, between March 2020 and February 2021, were used.

The association between time of admission and mortality was examined with multivariate logistic regression models.

**Results:** 3390 Covid-19 patients were included in the study, of which 1330 were women, the age was M(SD): 66.55(16.55) Death was reported for 451 patients. There was a significant decreasing trend in mortality by time of admission for the whole year with an OR: 0.86(0.77-0.96) p=0.005. No significant trend in mortality for women was observed OR: 1.00(0.85-1.19) p=0.959, while there was a significant decreasing trend for men OR: 0.78 (0.68-0.90) p=0.001

**Discussion:** The health policies put in place, the scientific evidence developed by researchers, and the experienced acquired by clinicians, are likely to explain this improvement in mortality. More epidemiological and clinical studies addressing trends of mortality in patients with different sociodemographic and clinical profile and the improvement of clinical outcomes are required. Future research may address the safety and efficacy of interventions specifically in female patients.

## Introduction

At the beginning of the pandemic caused by SARS-CoV-2 clinicians experienced a sudden increase in their workload. There was an insufficient number of beds available in hospital wards and intensive care units. The scientific evidence on the management of the infection was also very limited at the time. This all made difficult to provide an adequate healthcare. Since then, the health services have been reorganized, researchers have produced new evidence to help medical decisions, and clinicians have acquired experience on the management of the infection. The safety and efficacy of the health policy and clinical interventions is currently supported by different degrees of evidence.^1^ However, these interventions have in most cases been investigated one by one, and their results assessed after a short time for all patients with Covid-19. It is still poorly understood if the development of health services, clinical management, and scientific evidence is associated with better medical outcomes, sustained in the long term, for patients of each gender.^2-6^ Strong evidence on the trends of mortality would help to evaluate the progress made so far, and inform future clinical practice, research, and health policies. This study presents the trends in mortality associated with Covid-19 for women and men during the first year of the pandemic.

## Methods

This study was based in the 17 Spanish hospitals of the private healthcare provider HM Hospitales, placed in the provinces of Barcelona, Coruña, León, Madrid, Pontevedra, and Toledo.^7^ Sociodemographic, clinical, and mortality data from all patients with Covid-19, who had been discharged alive, or had died after being admitted, between March 2020 and February 2021, were used. Patients who were still admitted at the end of the study period, or had been transferred to another hospital, were not included. All patients had been diagnosed with polymerase chain reaction test of respiratory samples for SARS-CoV-2.

Time of admission was categorized in three-months period, March to May 2020, June to August 2020, September to November 2020, and December 2020 to February 2021. The association between time of admission and mortality was examined with logistic regression models, first univariate, then adjusted for age and gender, and finally also adjusted for saturation of oxygen on admission.^8^ This association was then examined separately for women and men. Sensitivity analysis was conducted with time of admission categorized in four-month periods, which made the number of individuals and events larger in each time category. The software Stata 11 was used for all the analysis.

## Results

A total of 3390 Covid-19 patients were included in the study, of which 1330 were women, the age was M(SD): 66.55(16.55) and the saturation of oxygen on admission was M(SD): 93% (4). The number of admissions in each three-months period varied, between 110 in June-August 2020 and 2060 in March-May 2057. Death was reported for 451 patients. The highest mortality rate was in 15.75% and it was recorded in the period March-May 2020. It was followed by the largest reduction in mortality, which went down to 8.18% in the following three-months period, June-August 2020 (Table 1 and Figure 1).

**Table 1.** Description of patients admitted throughout the study and in each three months period (* When estimates are presented separately for gender categories, adjustment was only for age in model 2 and for age and saturation of oxygen in model 3)

|  |  | All sample | Women | Men |
| --- | --- | --- | --- | --- |
| All sample | N | 3390 | 1330 | 2060 |
|  | Dead | 451(13.30) | 157(11.80) | 294(14.27) |
| Age M(SD) | n | 3390 | 1330 | 2060 |
|  | M(SD) | 66.55(16.55) | 68.69(17.19) | 65.17(15.97) |
| Saturation of Oxygen % | n | 2913 | 1109 | 1804 |
|  | M(SD) | 93 (4) | 94(4) | 93(4) |
| Admission in March-May 2020 | n | 2057 | 826 | 1232 |
|  | Dead | 324(15.75) | 107(12.95) | 217(17.63) |
| Admission in June-August 2020 | n | 110 | 42 | 68 |
|  | Dead | 9(8.18) | 3(7.14) | 6(8.82) |
| Admission in September-November 2020 | n | 513 | 188 | 325 |
|  | Dead | 48(9.36) | 16(8.51) | 32(9.85) |
| Admission in December 2020-February 2021 | n | 710 | 274 | 436 |
|  | Dead | 70(9.86) | 31(11.31) | 39(8.94) |
| Association of period of admission and death | Model 1 - Univariate analysis | 0.81 (0.74-0.88)<br>p<0.001 | 0.92 (0.80- 1.05)<br>p=0.215 | 0.75 (0.67-0.84)<br>p<0.001 |
|  | Model 2- Adjusted for age and gender * | 0.83 (0.76-0.92)<br>p<0.001 | 0.98 (0.85- 1.14)<br>p=0.800 | 0.75 (0.67-0.86)<br>p<0.001 |
|  | Model 3- Adjusted for age, gender and saturation of oxygen* | 0.86(0.77-0.96)<br>p=0.005 | 1.00(0.85-1.19)<br>p=0.959 | 0.78 (0.68-0.90)<br>p=0.001 |

**Figure 1.**
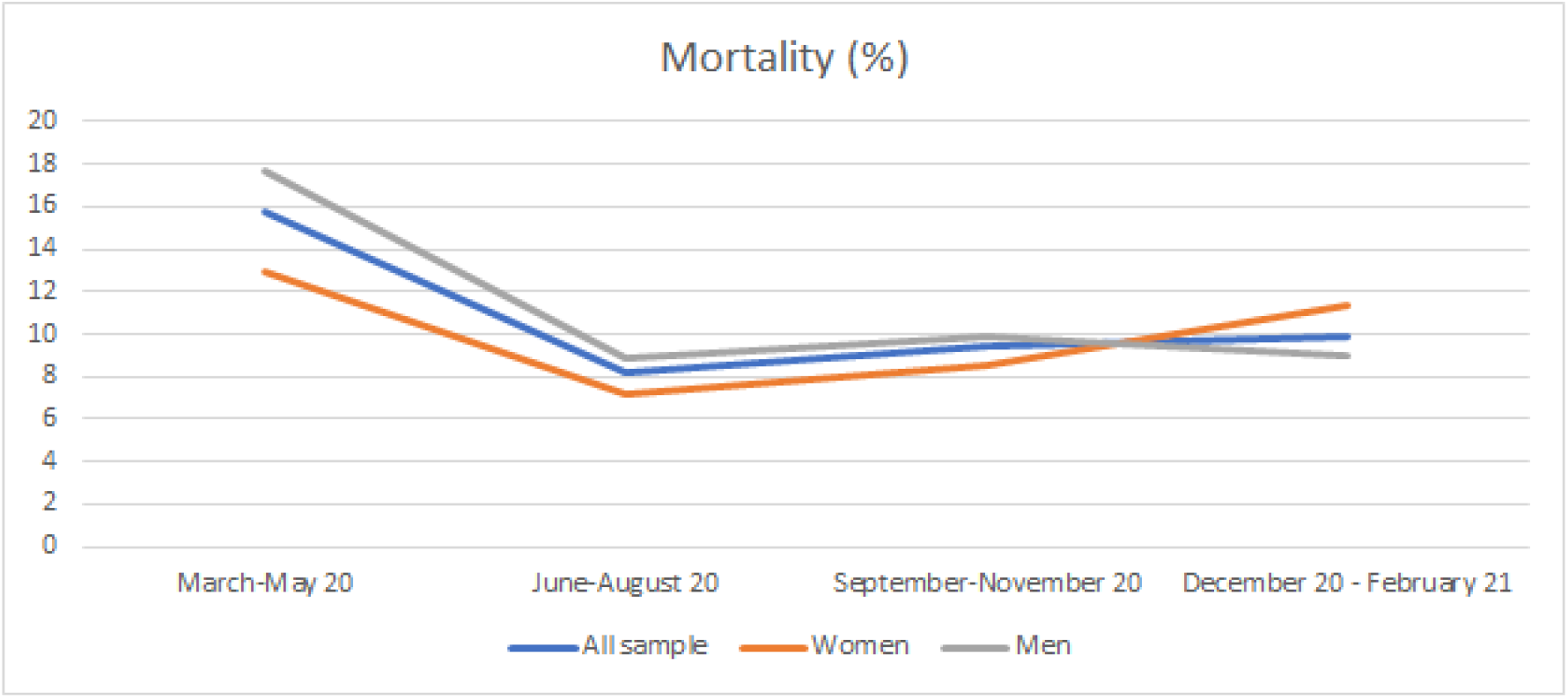
Mortality by three-month periods

There was a significant decreasing trend in mortality by time of admission for the whole year with an OR: 0.86(0.77-0.96) p=0.005. When the analysis was conducted for gender categories separately, no significant trend in mortality for women was observed OR: 1.00(0.85-1.19) p=0.959, while there was a significant decreasing trend for men OR: 0.78 (0.68-0.90) p=0.001 (Table 1) These trends were all consistent with the ones obtained when the analyses were conducted with time of admission categorized in four-month periods.

## Discussion

There was a significant decreasing trend in mortality among patients admitted with Covid-19 in the first year of the pandemic, with the largest reduction in deaths in the first few months. This decrease was significant for male, but not for female, patients.

The health policies put in place, the scientific evidence developed by researchers, and the experienced acquired by clinicians, are likely to explain this improvement in mortality. Treatments that mainly address the immune response to the infection, which is implicated in the higher mortality for men, may explain why the death rate in women does not improve.^1,9,10^ Behavioural changes among women, around the prevention of Covid-19, care for relatives, or seeking medical help, during the pandemic, may have increased their risk.^9^ It is also possible that the lower number of female patients included in the study and the reduced statistical power for the analysis, compared to the one available for male patients, may have not allowed to observe a decreasing trend in mortality.

This study has strengths and limitations. Patients were not randomized and the differences in mortality may be explained by factors other than the development of health services, clinical management, and scientific evidence. However, it should be noted that a randomized study investigating all these factors in different months would not be feasible. The observation of a large number of patients, all those discharged from 17 hospitals, the adjustment of statistical models for reported predictors of mortality,^8^ and the consistency of the results with the ones obtained in the sensitivity analysis, are strengths of this research.

This study provides encouraging results for policy makers, hospital clinicians, and researchers to continue their work. However, more epidemiological studies in larger databases looking at trends of mortality in patients with different sociodemographic and clinical profile are required. The death rate of Covid-19 patients remains high and more studies to improve clinical outcomes are also necessary. Future research may address the safety and efficacy of innovative or repurposed interventions specifically in female patients.

## Data Availability

Data requests should be addressed to the corresponding author

## Acknowledgements

Thank you to all clinicians and administrators of Hospitales HM (Spain) on which data this study is based.

## Ethical Approval

The Ethics Committe to HM Hospitales approved this study

